# Extensive validation and prospective observation of the impact of an AI-based rapid antibiotics susceptibility prediction platform in multiple medical centers

**DOI:** 10.1101/2021.06.23.21259391

**Authors:** Hsin-Yao Wang, Chia-Ru Chung, Yi-Ju Tseng, Jia-Ruei Yu, Chao-Jung Chen, Min-Hsien Wu, Ting-Wei Lin, Wan-Ting Huang, Tsui-Ping Liu, Tzong-Yi Lee, Jorng-Tzong Horng, Jang-Jih Lu

## Abstract

**Importance:** No medical artificial intelligence (AI) has been robustly validated and deployed in a clinical laboratory in real-world settings, and the clinical impact of the medical AI remains unknown.

**Objective:** To deploy a medical AI platform for rapid antibiotics susceptibility test (AST) prediction, and evaluate its clinical impacts.

**Design:** A medical AI platform, XBugHunter, was extensively validated (internal validation, time-wise validation, and independent testing) with data between May 22, 2013 and June 30, 2019. The clinical impact was evaluated based on a prospective observation from February 1 to September 30, 2020 during deployment.

**Setting:** Data was collected in two tertiary medical centers in Taiwan, and the AI was deployed in a tertiary medical center.

**Participants:** For validation, 90,064 consecutive cases were included. During the deployment, a prospective observational cohort of 1,490 consecutive cases was collected.

**Exposures:** AST prediction from XBugHunter

**Main outcomes and Measures:** Diagnostic performance of XBugHunter was evaluated during validation. The clinical impact was evaluated in terms of the saving of inappropriate antibiotics prescription, AST turn-around-time, and mortality of bacteremia during deployment.

**Results:** Predictive models consistently performed well in the extensive validations. In the deployment, XBugHunter’s predictive sensitivity and specificity for *Staphylococcus aureus* (oxacillin) were 0.95 (95% CI, 0.82–0.98) and 0.97 (95% CI, 0.94–0.99), respectively. For *Acinetobacter baumannii* (multiple drugs), the sensitivity was 0.95 (95% CI, 0.91–0.99) and specificity was 0.93 (95% CI, 0.88–0.98). The turn-around-time reduction on reporting AST of blood cultures was 35.72 h (standard deviation: 15.55 h). Death within 28 days occurred in 28 of 162 *S. aureus* bacteremia patients (17.28%) in the XBugHunter intervention group, which was lower than the 28 days’ mortality rate (28.06% [55/196]) in the same period of time in 2019, without XBugHunter. The relative risk reduction was 38.4% (relative risk, 0.62; 95% CI, 0.41–0.92). Regarding antibiotic prescriptions, 2723.7 defined daily dose per year of inappropriate antibiotics could be avoided for treating *S. aureus* by deploying XBugHunter.

**Conclusions and Relevance:** Among *S. aureus* bacteremia patients, this study demonstrated that XBugHunter can prevent inappropriate antibiotic use, and the adjustment of antibiotic treatment can yield lower mortality.

**Key Points:** *Question:* What is the clinical impact of XBugHunter, a machine learning-based, antibiotic susceptibility test prediction platform?

*Findings:* In the prospective observational cohort of deploying XBugHunter, the reduction of turn-around-time of reporting antibiotic susceptibility test was 35.72h. The reduction of *S. aureus* bacteremia mortality rate was 10.78%, and the estimated saving of inappropriate antibiotics uses was 2723.7 defined daily dose per year.

*Meaning:* Deployment of XBugHunter provides a more rapid report of antibiotic susceptibility test, and thus reduces inappropriate antibiotics prescription and mortality of *S. aureus* bacteremia.

## Introduction

Infectious diseases are responsible for a considerable healthcare burden in the current healthcare system. This burden has continued to increase in recent years. It is estimated that infectious diseases would cause more deaths than cancers by 2050.^1^ The timely and accurate antibiotic susceptibility test (AST) is the key to the successful treatment of bacterial infections.^2^ Moreover, rapid AST would guide correct antibiotics use in the early stage of infectious diseases,^3^ reduce mortality of infectious diseases,^2^ and prevent development of resistance due to inappropriate antibiotics use.^4^ Rapid AST is also reported to influence empiric antibiotic selections for bacteremia to facilitate early de-escalation of therapy without compromising the adequacy of antibiotic coverage.^3^ In brief, rapid and accurate AST plays a central role in the treatment of individual infection and surveillance of antimicrobial use. In the past decades, resistant pathogens, including *Enterococcus faecium, Staphylococcus aureus, Klebsiella pneumoniae, Acinetobacter baumannii, Pseudomonas aeruginosa*, and *Enterobacter spp*. (ESKAPE) have posed considerable threats to global health.^5^ Even though the correct treatment of ESKAPE infection largely depends on AST, typical AST methods used in the current clinical microbiology laboratories cost in terms of the long turn-around-time (TAT).^6^ The long TAT of AST exposes infectious diseases under long empiric antibiotic use. Hence, developing a rapid and accurate AST for guiding correct antibiotic use is of clinical interest.

Matrix-assisted laser desorption ionization time-of-flight (MALDI-TOF) mass spectrometry (MS) is a relatively novel method with a broad applicability range in clinical microbiology laboratories.^7^ MALDI-TOF MS is typically used as a tool for the identification of bacterial species.^7^ Additionally, MALDI-TOF MS holds promise for rapid reporting of other bacterial characteristics, including strain typing^8-11^ and AST.^6,12,13^ Rapid AST based on MALDI-TOF spectra reduces TAT of AST by more than 30 h,^14^ and requires only spectra generated in routine workflow.^6^ This means that no additional laboratory analysis is necessary. Thus, providing rapid AST from routine MALDI-TOF spectra is promising because it is potentially cost-effective and easy to integrate into current workflow of clinical microbiology laboratory.^6,14,15^ Based on the approach, accurate prediction of oxacillin resistance for *Staphylococcus aureus* has been reported by two independent teams.^6,14^ MALDI-TOF spectra-based AST for other species of ESKAPE, such as *Enterococcus faecium, Klebsiella pneumoniae, Pseudomonas aeruginosa*, has also been reported recently.^14,15^ In these studies, machine learning (ML) algorithms were adopted for the analysis of the complex data of MS spectra.^6,14,15^ Meanwhile, large data were used for training and validating the ML models.^6,14,15^ However, application of the MALDI-TOF spectra-based rapid AST has not yet been supported by adequate evidence from its implementation in real-world cases.^16^ Individualized clinical ML models offer the promise of both reducing broad-spectrum antibiotic use and preserving/improving adequacy of treatment, but few have been validated in the clinical setting.^3^ In addition, high-predictive accuracy could be achieved only in a small part (<50%) of the bacterial strains.^14^ The approach of MALDI-TOF spectra-based rapid AST has not been developed extensively for the important pathogens (e.g., ESKAPE). All these unfavorable factors still hinder the widespread use of MALDI-TOF spectra-based AST.

Similar to typical diagnostic markers, digital ML models also require validation in clinical studies to show the impact on relevant outcomes.^7^ Herein, we trained, validated, independently tested, and implemented a platform (XBugHunter) capable of reporting MALDI-TOF spectra-based AST for ESKAPE. In addition to the prediction of AST, we also implemented and evaluated XBugHunter’s impact on the TAT of AST, which provided guidance for antibiotic use and survival rate of bacteremia in a prospective observational cohort.

## Materials and methods

### Data source for training and independently validation of XBugHunter

The study design is shown in Figure 1. Data used for training and validation of XBugHunter were collected consecutively from the clinical microbiology laboratories of two tertiary medical centers in Taiwan (Chang Gung Memorial Hospital (CGMH) Linkou branch and Kaohsiung branch) between May 22, 2013, and June 30, 2019. Clinical microbiology laboratories conducted all the routine microbiological tests ordered from the hospitals. Six superbugs (ESKAPE) with the corresponding important antibiotics were included in XBugHunter. In total, 90,064 cases were included for training and validating models of XBugHunter. Of these cases, 74,722 cases were collected from the CGMH Linkou branch, while 15,342 cases were collected from the CGMH Kaohsiung branch. Detailed information regarding the pathogens and their antibiotics is illustrated in Figure 2. The study was approved by the Institutional Review Board of the Chang Gung Medical Foundation (No. 201900767B0). Standards for Reporting of Diagnostic Accuracy 2015^17^ and the Transparent Reporting of a Multivariable Prediction Model for Individual Prognosis or Diagnosis reporting guidelines^18^ were followed.

**Figure 1.**
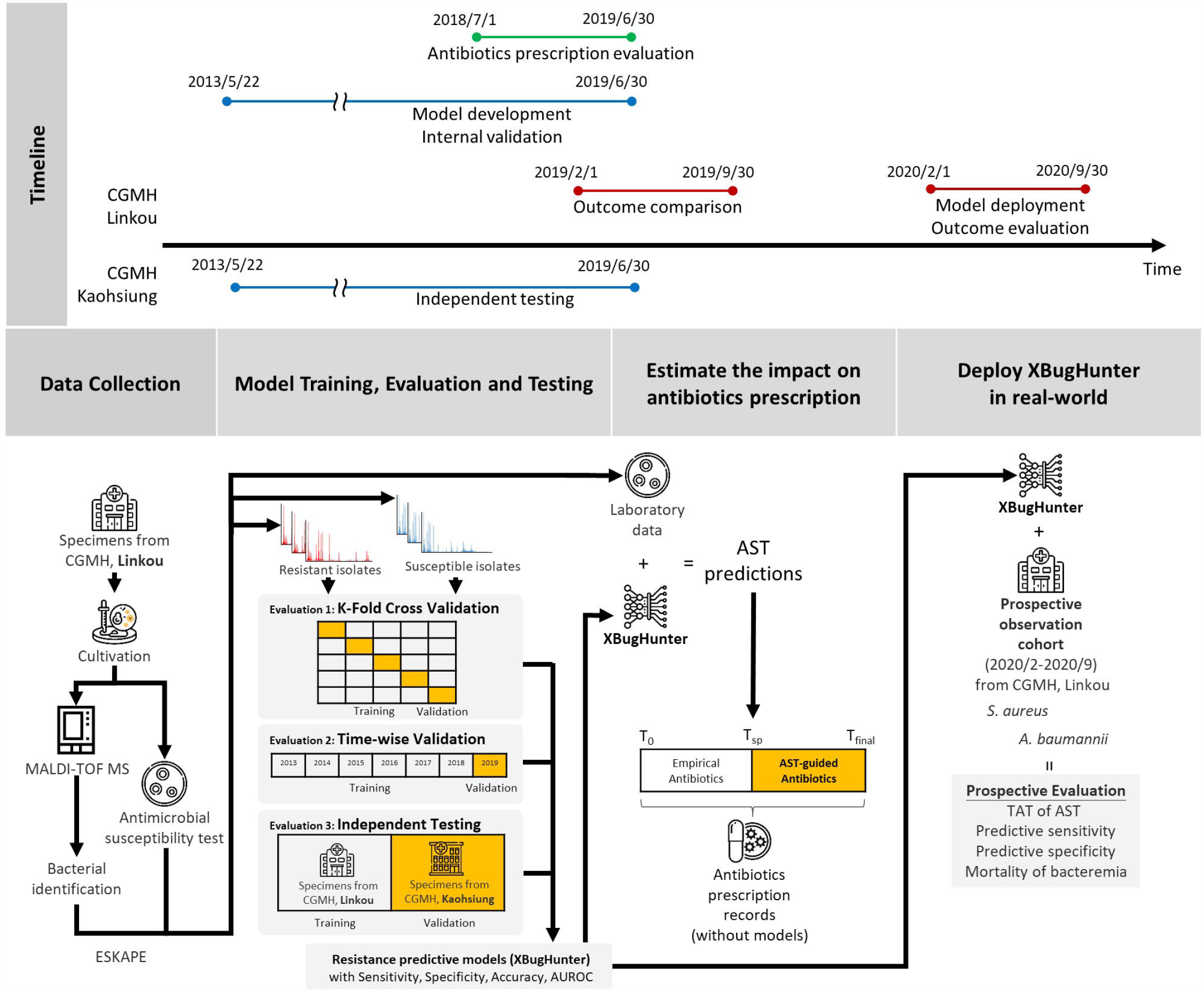
Scheme of the study. Major measurements in this study include the predictive performance of XBugHunter in internal validation, time-wise validation, and independent testing. The impact of antibiotic prescription was also estimated. Moreover, the predictive performance of deploying XBugHunter in a clinical setting was evaluated in terms of the clinical sensitivity/specificity of predicting resistant strains. The mortality of *S. aureus* bacteremia was compared between the XBugHunter-intervention group in the prospective observation cohort and the non-XBugHunter-intervention group (CGMH, Chang Gung Memorial Hospital; ESKAPE, *Enterococcus faecium, Staphylococcus aureus, Klebsiella pneumoniae, Acinetobacter baumannii, Pseudomonas aeruginosa*, and *Enterobacter spp*.; AST: antibiotic susceptibility test; T_0_: time of specimen collection; T_sp_: time of reporting bacterial species; T_final_: time of reporting AST).

**Figure 2.**
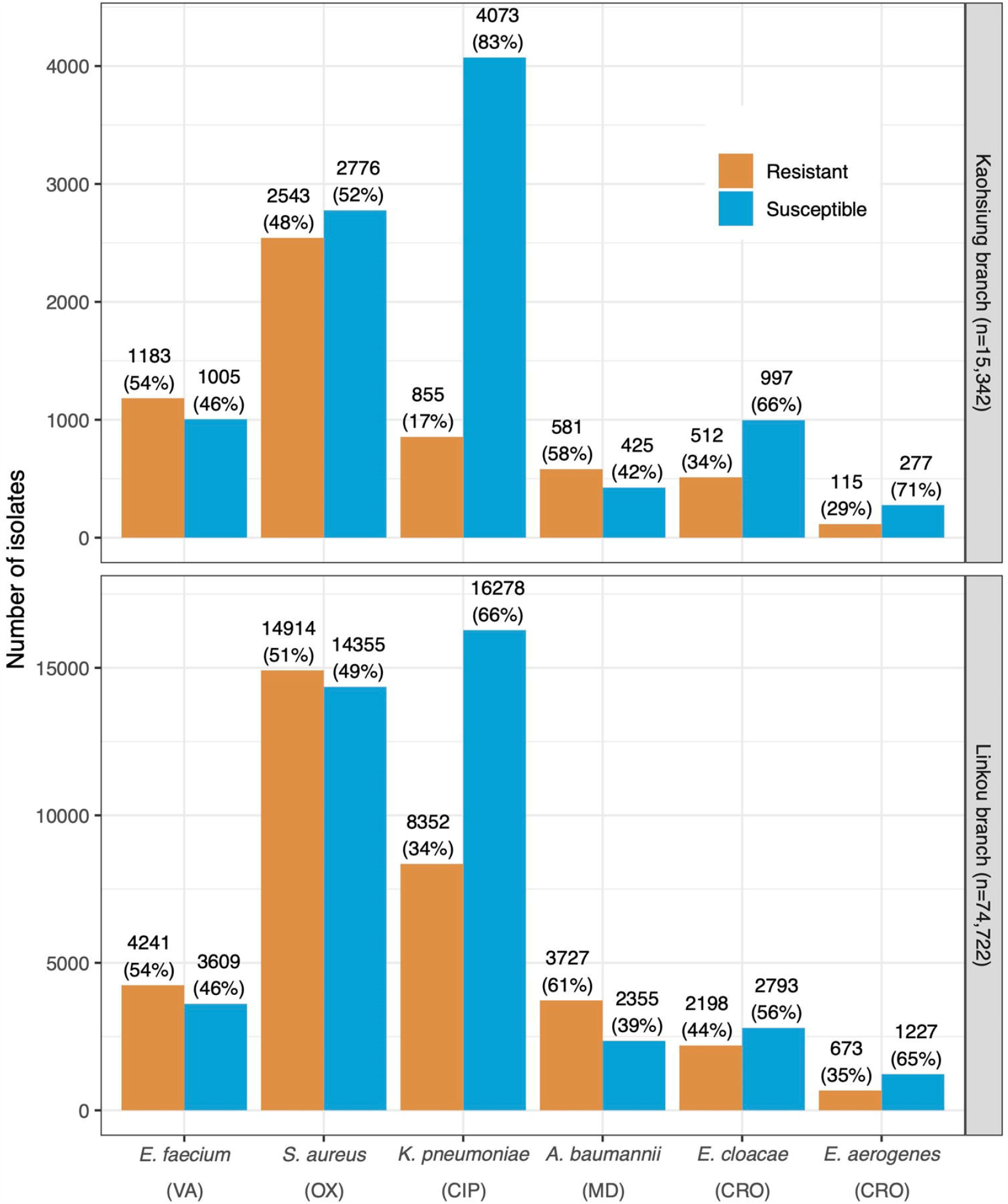
Distribution of susceptible and resistant ESKAPE isolates in Linkou Chang Gung Memorial Hospital (CGMH) and Kaohsiung CGMH. (VA: vancomycin; OX: oxacillin; CIP: ciprofloxacin; MD: multiple drugs; CRO: ceftriaxone).

### Identification of bacterial species and antibiotic susceptibility test

Bacterial species were identified using MALDI-TOF spectra acquired with the use of a Microflex LT mass spectrometer and analyzed using Biotyper 3.1 (Bruker Daltonik GmbH, Bremen, Germany). The details of bacterial species identification and AST are provided in the **Supplemental Materials**.

### Preprocessing of MALDI-TOF MS spectra prior to the analysis by XBugHunter

In this study, we adopted MALDIquant, an R packages, to preprocess all the MS spectra.^19^ Specifically, the top-hat method was implemented to deal with the baseline correction. The noise of the MS spectrum was detected based on the calculation of the median absolute deviation with a half-window size of 10. The threshold of the signal-to-noise ratio was 5. After preprocessing the raw signals, we created the “type templates” for each species based on the incidence of specific peaks in their MALDI-TOF MS spectra to handle the issue of peak shifting.^9,12^ Further, these processed MS spectra were used to construct predictive models on XBugHunter.

### Feature investigation and model development and validation

In this study, the random forest (RF) was adopted to investigate the feature importance. Specifically, the feature importance was determined based on the Gini importance, as provided by Python in the scikit-learn package^20^. Note that the number of trees in the RF was 1000, and the nodes were expanded until all leaves were pure or contained less than 1. After the selection of informative MS peaks, various ML-based methods, such as decision tree (DT), logistic regression (LR), RF, support vector machines (SVM), and artificial neural networks (NN), were employed to construct the prediction models. Specifically, 10-fold cross-validation was used to determine the possible parameters. Note that these methods were implemented with the use of the scikit-learn package provided in Python^20^. Details are described in the **Supplemental Materials**.

### Implementation of XBugHunter in a prospective observational cohort and evaluation of the clinical impacts on TAT and survival of bacteremia

We deployed XBugHunter in the routine practice of clinical microbiology laboratories in the CGMH Linkou branch on February 1, 2020. We acquired the data of the prospective observational cohort between February 1, 2020 and September 30, 2020 for analysis. During deployment, *S. aureus* (oxacillin) and *A. baumannii* (multiple drugs [MD]) models of XBugHunter were used to report the predictive AST. We uploaded the whole-cell MALDI-TOF MS spectra in XBugHunter (Supplemental Video) immediately after the spectra were generated for the identification of bacterial species. Clinical pathologists provided predictive ASTs for eligible cases based on the predictions of XBugHunter. The gray zone method was adopted to define the eligible cases to report the AST predictions. The gray zone (θ) was determined to be approximately 0.5. The cases whose predictive probabilities were located between 0.5 - θ and 0.5 + θ were considered to have insufficient confidence, and the predictive AST was not reported to the clinical physicians (Supplemental eFigure 1). Based on a prospective observational cohort, we evaluated the impact of XBugHunter on the TAT of ASTs by comparing the TAT of the predictive ASTs and the TAT of typical ASTs. Moreover, the impact of XBugHunter on the survival of *S. aureus* bacteremia was compared with that of *S. aureus* bacteremia recorded between February 1, 2019 and September 30, 2019.

### Impact on antibiotic prescription by XBugHunter

We evaluated the impact of XBugHunter on antibiotic prescription with the use of a dataset collected from the CGMH Linkou branch between July 1, 2018 and June 30, 2019. The time points for bacterial cultures and antibiotic prescriptions were recorded in the dataset. During this period, XBugHunter was not implemented in the clinical setting. We evaluated the impact on antibiotic prescription for treating *S. aureus* infections by simulating using XBugHunter in the period of time (July 1, 2018 and June 30, 2019). XBugHunter was expected to provide predictive AST at the time of reporting the bacterial species (T_sp_) (Figure 1). AST was conducted according to the bacterial species and provided days later (T_final_). We evaluated the amount of antibiotics that could be corrected by XBugHunter between T_sp_ and T_final_.

### Identification of protein markers

The details are provided in the **Supplemental Materials**.

## Results

### Predictive performance on internal validation, independent testing

We used 74,722 cases from Linkou CGMH for internal validation and 15,342 cases from Kaohsiung CGMH for independent testing. The resistant-to-susceptible ratios were comparable between the Linkou CGMH and Kaohsiung CGMH (Figure 2). In the internal validation based on the 74,722 cases, the predictive models typically yielded areas under the receiver operating characteristic curves (AUROC) higher than 0.80, except *for K. pneumoniae* (ciprofloxacin), *E. aerogenes* (ceftriaxone), and *E. cloacae* (ceftriaxone) models (Figure 3). Specifically, the AUROC was 0.9406 (95% confidence interval [CI], 0.9380– 0.9433) for *S. aureus* (oxacillin), 0.9515 (95% CI, 0.9473–0.9554) for *A. baumannii* (MD), 0.8294 (95% CI, 0.8203–0.8383) for *E. faecium* (vancomycin), 0.7993 (95% CI, 0.7936– 0.8050) for *K. pneumoniae* (ciprofloxacin), 0.6600 (95% CI, 0.6343–0.6857) for *E. aerogenes* (ceftriaxone), and 0.7386 (95% CI, 0.7247–0.7523) for *E. cloacae* (ceftriaxone) (Figure 3(A)). In the independent testing using data from Kaohsiung CGMH, the AUROCs were similar to those obtained in the internal validation (Figure 3(B)). Likewise, in the time-wise independent testing (Figure 3(D)), the predictive performances were not inferior to the performances in the time-wise internal validation (Figure 3(C)). Briefly, in both the location-wise independent testing and time-wise independent testing, XBugHunter’s predictive performance was robust. In the ML models, the performances of *S. aureus* (oxacillin) and *A. baumannii* (MD) models were consistent in the comprehensive validations and superior to those of other models of pathogens. In addition, *S. aureus* (oxacillin) and *A. baumannii* (MD) models also performed consistently for different types of specimens in both internal validation (Figures 4(A) and 4(C)) and independent testing (Figures 4(B) and 4(D)).

**Figure 3.**
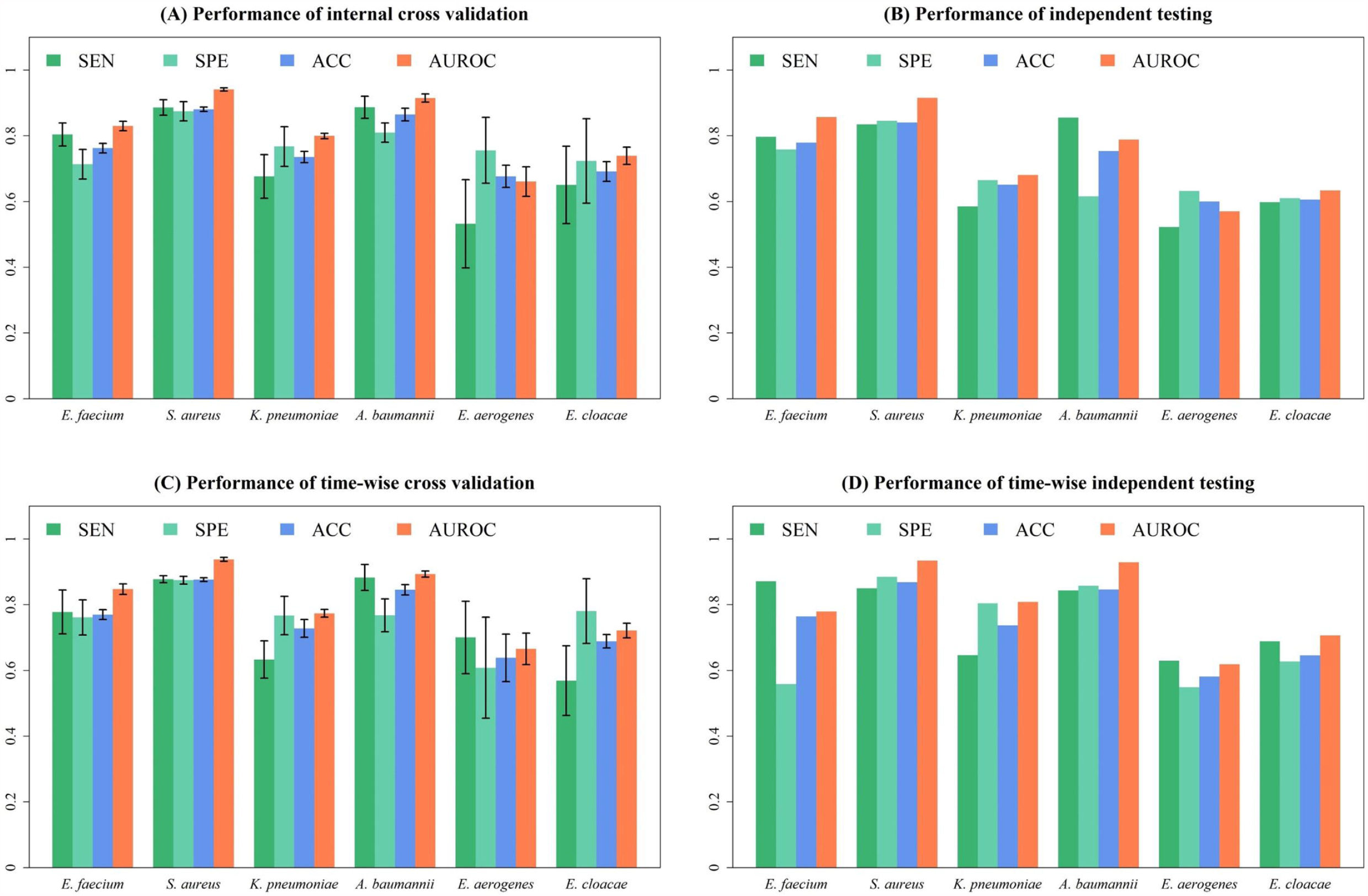
Predictive performance for ESKAPE in the comprehensive validations. The error bars indicate the 95% confidence intervals of the predictive performance. SEN: sensitivity; SPE: specificity; ACC: accuracy; AUROC: area under receiver operating characteristic curve.

**Figure 4.**
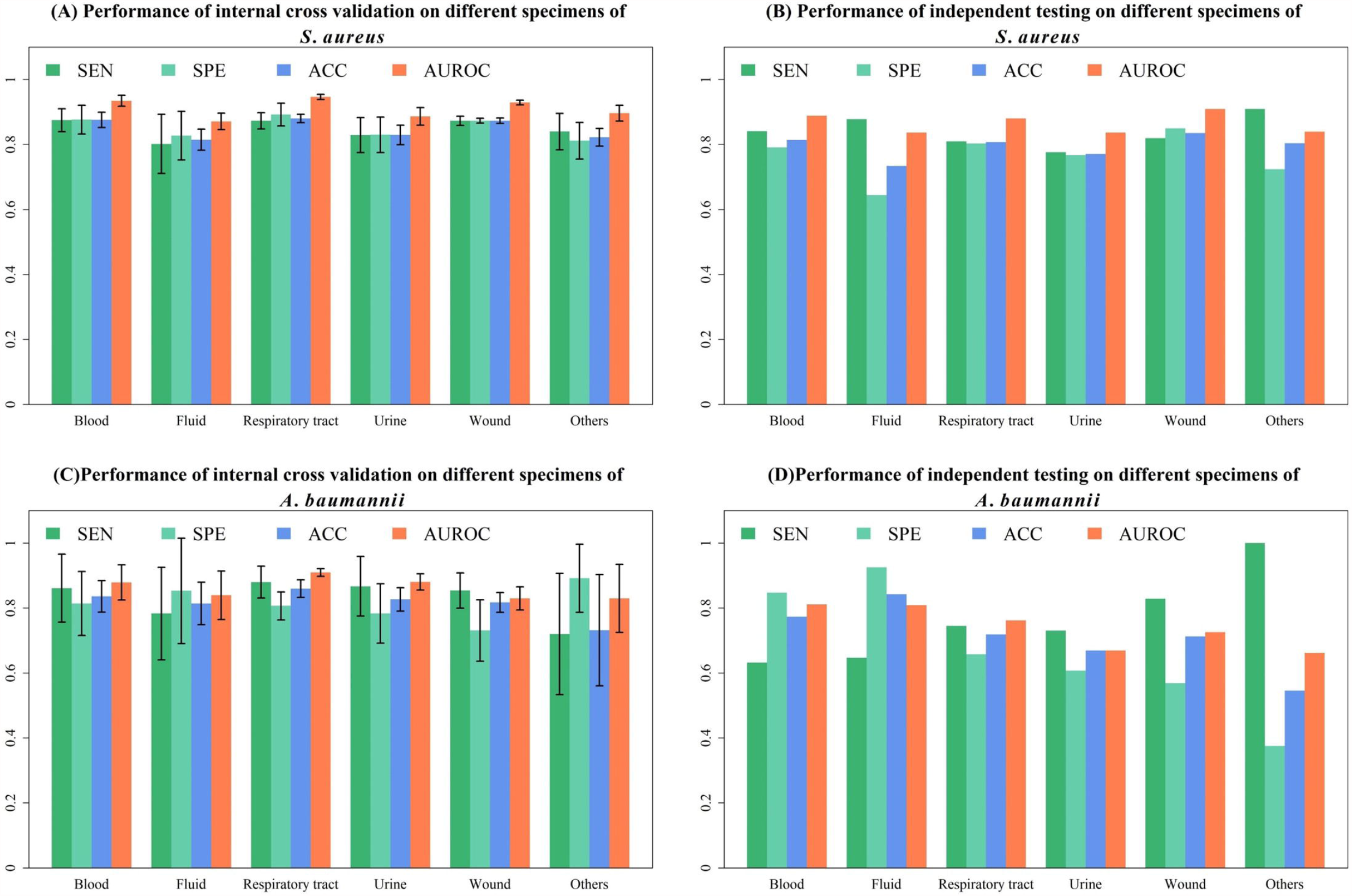
Predictive performance for *S. aureus* (oxacillin) and *A. baumannii* (multiple drugs) in different specimen types in internal validation and independent testing. The error bars indicate the 95% confidence intervals of the predictive performance. SEN: sensitivity; SPE: specificity; ACC: accuracy; AUROC: area under receiver operating characteristic curve.

### Implement XBugHunter in clinical settings and estimate the impacts on reduction of AST TAT and on mortality of *S. aureus* bacteremia

Based on internal validation, independent testing, and time-wise independent testing, the performance of XBugHunter was considered stable. We implemented *S. aureus* (oxacillin) and *A. baumannii* (multiple drug) models of XBugHunter in a clinical setting and conducted evaluations on a prospective observational cohort. A total of 1,490 consecutive cases (*S. aureus* and *A. baumannii*) was included, in which there were 211 bacteremia cases. For *the S. aureus* (oxacillin) model, the sensitivity was 0.9515 (95% CI, 0.8232–0.9797) and specificity was 0.9667 (95% CI, 0.9431–0.9902). For *the A. baumannii* (multiple drug) model, the sensitivity was 0.9518 (95% CI, 0.9094–0.9942) and specificity was 0.9333 (95% CI, 0.8839–0.9827). When XBugHunter was deployed, the TAT reduction on blood cultures used for diagnosing *S. aureus* bacteremia was 35.72 h (standard deviation: 15.55 h).

In addition to TAT reduction on AST, the clinical impact of XBugHunter was also estimated. In the prospective observational cohort using XBugHunter, the mortality rate of *S. aureus* bacteremia was 17.28% (28/162); in the same period of time in 2019, the mortality rate of *S. aureus* bacteremia was 28.06% (55/196). The absolute risk reduction on mortality of *S. aureus* bacteremia was 10.78%, whereas the relative risk reduction was 38.4% (relative risk, 0.62; 95% CI, 0.41–0.92)

### Reducing inappropriate antibiotic prescriptions

In addition to reducing the TAT of AST and reducing the mortality of bacteremia, we also estimated the potential impact of XBugHunter on antibiotic surveillance. Based on the retrospective dataset, we simulated the impact of deploying XBugHunter on *S. aureus* infections. For the treatment of *S. aureus* infections, empirical antibiotics were generally divided into beta-lactams, glycopeptides, and other antibiotics. Glycopeptides are appropriate, and beta-lactams are inappropriate for treating MRSA infections. In contrast, for MSSA cases, beta-lactams are considered appropriate, whereas glycopeptides are considered inappropriate. In the one-year simulation period, the predictive AST of XBugHunter prevented inappropriate use of 1665.8 defined daily dose (DDD) of antibiotic for treating MRSA (Supplemental eTable 2(a)), and that of 1057.9 DDD of antibiotics for treating MSSA (Supplemental eTable 2(b)). In total, inappropriate use of 2723.7 DDD of antibiotics could be avoided by deploying XBugHunter. The average saving of inappropriate antibiotic uses was estimated to be 1.57 DDD per predictive AST provided by XBugHunter.

### Informative peaks for predicting AST

The results are provided in the **Supplemental Materials**.

## Discussion

We developed, independently tested, and implemented a ML-based platform (XBugHunter) that can provide rapid AST for ESKAPE based on the MALDI-TOF spectrum. TAT of AST could be accelerated by approximately 36 h for bacteremia. Based on the rapid AST from XBugHunter, the accuracy of antibiotic prescription could be improved, and unnecessary antibiotic use could be avoided. In the prospective observational cohort, 28 days mortality rate of *S. aureus* bacteremia was reduced according to the rapid AST from XBugHunter. XBugHunter harnessed ML in analyzing MALDI-TOF spectra and would have impacts on managing ESKAPE infections in clinical settings in the real world.

In the current technological era, tremendous interest has been generated in the development of artificial intelligence (AI) or ML models for applications in the medical field. However, only a few medical AI models have been extensively validated and deployed in clinical practice.^21^ The gap between the development of medical AI models and the deployment of AI is obvious. Generally, integrating larger amounts of data (laboratory, clinical, geographical) is needed to validate a medical AI.^22^ Specifically, external validation or independent testing for a medical AI estimates the robustness of a medical AI in a variety of institutes. To examine the robustness of XBugHunter, we conducted internal validation (Figure 3(A) and 3(C)), independent testing (Figure 3(B)), time-wise independent testing (Figure 3(D)), amid a various of ML algorithms (Supplemental eFigure 3), and specimen-wise validation (Supplemental eFigure 4). The steady performance among the validations indicated that XBugHunter can perform well in different institutes (independent testing, Figure 3(B)) and different periods of time (time-wise independent testing, Figure 3(D)). On this basis, we deployed XBugHunter in our routine practice to predict susceptibility results for *S. aureus* and *A. baumannii*. The implementation of the medical AI also showed favorable results (accuracy higher than 0.95) for actual application in clinical settings.

Another gap in the deployment of medical AI is the lack of evidence or guidance for its application in addition to its capacity to report predictive performance. Few physicians during the era received training in quantitative methods and physicians generally trusted their own subjective intuition more than the empirical output of an algorithm.^21^ To make a medical AI understood to clinical physicians, clinical impacts would be more convincing. In this study, we reported evidence of clinical impact in addition to predictive performance. The time to the appropriate antibiotic administration is one of the keys in the management of infectious diseases.^2^ Appropriate antibiotic administrations within 48 h are associated with reduced mortality in bacteremia patients.^2^ Appropriate antibiotic administrations depend largely on *in vitro* testing of AST. The advantageous characteristics of XBugHunter include rapidness and appropriate accuracy (Figures 3 and 4), especially when modified with the gray zone method. Moreover, XBugHunter could save 1.57 DDD unnecessary antibiotic prescriptions per prediction in treating *S. aureus* infections. In the deployment, a lower risk of death was observed within 28 days in the XBugHunter-guided group compared with the non-XBugHunter-guided group among *S. aureus* bacteremia patients. The real-world evidence of XBugHunter would be a key to overcoming the gap between development and real application in clinical practice.

Explanation of medical AIs is another weakness that may mitigate the clinical confidence of physicians in medical AI. In this study, we evaluated the most important features that were used in the predictive models (Supplemental eTable 1). Furthermore, we visualized the important features in the spectrum for each prediction so that users could easily understand the reasons for each predictive result (Supplemental Video). On this basis, we investigated further the identity of the important features. In the spectrum, the important features are essentially peptides. We identified that *m/z* 3006 is a delta-lysin family PSM, which is in agreement with previous studies.^23,24^ Besides *m/z* 3006, still there are tens of important peaks that would be interesting for investigation of resistant mechanisms (Supplemental eTable 1). However, identification of the important peaks is labor-intensive, thus requiring the efforts of the entire research community.

This study was associated with several limitations. First, although we used the largest up-to-date real-world MALDI-TOF data, all the data were collected from different referral medical centers in Taiwan. The performance of XBugHunter when deployed in different areas or countries is not known. A cross-national study of XBugHunter deployment is required. Given the high diversity of microorganisms across areas/countries, it would be not possible that the current version of XBugHunter can be used in other areas/countries without adjustment. Training and validating ML models based on locally relevant MALDI-TOF data are favorable. Second, we adopted a gray-zone method for the ML models in the deployment of the XBugHunter in the clinical routine. The methodology enhanced the predictive performance (accuracy > 0.95); however, it excluded some of the cases that could not obtain adequate confidence in the prediction. The excluded portion of the gray zone may consist of rare strains. We plan to explore the constitution of the gray zone in future work. Finally, the microorganisms continue to evolve. It is not known how frequently ML models should be updated. The predictive performance at the time of deployment was stable. The change in predictive performance in long-term deployment is worthy of additional investigations.

## Conclusions

We developed, internally validated, independently testing, and deployed a MALDI-TOF-based medical AI (XBugHunter) to achieve a more rapid AST report. Deployment of XBugHunter could reduce inappropriate antibiotic prescription and reduce mortality in patients with *S. aureus* bacteremia.

## Data Availability

Data are available for individuals that are approved.

## Figure Legends

**Video**

**Title:** Demonstration of executing XBugHunter.

**Caption:** This video demonstrates executing XBugHunter in real world settings. Users can acquire rapid AST predictions by uploading whole-cell MALDI-TOF MS spectra on the user interface. In addition to predictive results, complete spectrum is visualized, and important features for prediction are highlighted. AST, Antibiotics susceptibility test; MALDI-TOF, Matrix-assisted laser desorption ionization time-of-flight; MS, mass spectrometry.

## Conflicts of Interest Disclosures

The authors have no affiliations with or involvement in any organization or entity with any financial interest or nonfinancial interest in the subject matter or materials discussed in this manuscript.

## Funding/Support

This work was supported by Chang Gung Memorial Hospital (CMRPG3L0401, CMRPG3L0431)

## Authors’ Contributions

Dr Wang had full access to all of the data in the study and takes responsibility for the integrity of the data and the accuracy of the data analysis.

Concept and design: Wang, Chung, Tseng.

Acquisition, analysis, or interpretation of data: Wang, Chung, Chen, Tseng, Lee, Horng, Lu. Drafting of the manuscript: Wang, Chung, Tseng, Yu.

Critical revision of the manuscript for important intellectual content: Wang, Chung, Tseng, Yu, Wu, Lin.

Statistical analysis: Chung, Tseng, Yu. Obtained funding: Horng, Lu.

Administrative, technical, or material support: Wu, Lin, Huang, Liu.

Supervision: Lee, Horng, Lu. Drs Lee, Horng, and Lu contributed equally to this work.

## Additional Contributions

We acknowledge the contribution of the patients and clinical staffs for the cohort included in this study.

## Notes

### Competing Interest Statement

The authors have declared no competing interest.

### Author Declarations

The study was approved by the Institutional Review Board of the Chang Gung Medical Foundation (No. 201900767B0)

