## Supplement materials for "Extensive validation and prospective observation of the impact of an AI-based rapid antibiotics susceptibility prediction platform in multiple medical centers"

eMethods

eTable 1. Top 15 RF importance of MS peaks.

eTable 2(a). XBugHunter’s potential reduction on anti-MRSA agents in a tertiary medical center for one year.

eTable 2(b). XBugHunter’s potential reduction on anti-MSSA agents in a tertiary medical center for one year.

eFigure 1. Adopting gray zone method to enhance predictive performance.

eFigure 2. Peptide sequence of *m/z* 3006.

eFigure 3(a). Internal validation by using different ML algorithms.

eFigure 3(b). Independent testing by using different ML algorithms.

eFigure 3(c). Time-wise independent testing by using different ML algorithms.

eFigure 4(a). Internal validation of different *S. aureus* ML models in a various of specimen types.

eFigure 4(b). Independent testing of different *S. aureus* ML models in a various of specimen types.

eFigure 4(c). Internal validation of different *A. baumannii* ML models in a various of specimen types.

eFigure 4(d). Independent testing of different *A. baumannii* ML models in a various of specimen types.

eReferences

This supplementary material has been provided by the authors to give readers additional information about their work.

### eMethods

#### Specimen processing, bacterial species identification, and antibiotics susceptibility tests

We received clinical specimens from all the wards continuously. The specimen types included blood, respiratory tract specimen (*ie,* sputum, bronchial wash, and bronchoalveolar lavage), sterile cavity fluid (*ie*, ascites, pleural effusion, pericardial effusion, cerebrospinal fluid, and synovial fluid), urine, and wound. Blood samples were collected from the patients in whom bacteremia was suspected, and cultured in trypticase soy broth (Becton Dickinson, MD, USA). Subculture culture was done for the positive blood culture bottles onto blood plate (BP) agar (Becton Dickinson, MD, USA). For sputum specimens, BP agar (Becton Dickinson), eosin methylene blue (EMB) agar (Becton Dickinson), CNA agar (Becton Dickinson), and chocolate agar (Becton Dickinson) were used for cultivation. For test of sterile cavity fluid, thioglycollate broth (Becton Dickinson) was used addition to BP, EMB, CNA, and chocolate agars. Urine specimens were quantitatively inoculated on BP and EMB agars. Saline (0.9%) was used for rinsing swabs that were used for collection of wound specimens, and then inoculated on BP, EMB, CNA, and chocolate agars. Pus specimens were inoculated in thioglycollate broth and on agars. We conducted the cultures in a CO_2_ incubator at 37℃ for 18–24 hours. Single colonies that recovered from the agars were used for species identification and antibiotics susceptibility tests. Bacterial species were identified by using matrix-assisted laser desorption ionization time-of-flight (MALDI-TOF) spectra (Bruker Daltonik GmbH, Bremen, Germany). Antibiotics susceptibilities were tested by using both microdilution (for blood and sterile cavity fluids specimens) and paper disc methods (other specimens). M50 instrument (Becton Dickinson) was used for testing minimal inhibitory concertation of antibiotics via microdilution method. The susceptibility of antibiotics was interpreted according to CLSI M100 of the corresponding years.

#### Development and validation of AST prediction models on XBugHunter

After the preprocessing of the MS and AST, we adopted several machine learning-based methods, decision tree (DT), logistic regression (LR), random forest (RF), support vector machine (SVM), and artificial neural network (NN), to construct the prediction models. More specifically, the 10-fold cross validation was used to determine the possible parameters. Note that these methods were implemented by scikit-learn package  provided in python^1^. We calculated four common metrics, sensitivity (SEN), specificity (SPE), accuracy (ACC), and area under the receiver operating characteristic curve (AUROC), to evaluate the prediction models. Except AUROC, their definitions are given below:

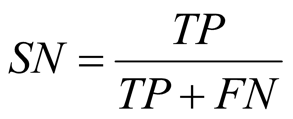

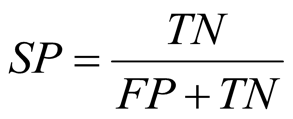

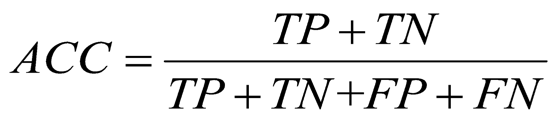
,

where TP means true positive and refers to the number of non-susceptible MS that were correctly predicted by the classifier, TN means true negatives and refers to the number of susceptible MS that were correctly predicted by the classifier, FP means false positives and refers to the number of non-susceptible MS that were incorrectly predicted by the classifier, and FN means false negative and refers to the number of susceptible MS that were incorrectly predicted by the classifier. In this study, the model with the highest AUROC was regarded as the best model and was further provided to evaluate the clinical efficacy.

#### Feature importance for the predictive models

In this study, we adopted the random forest to investigate the feature importance. Specifically, the importance of a feature is determined by Gini importance which is provided by python in scikit-learn package^1^. Note that the number of trees in the random forest was 1000 and the nodes were expanded until all leaves were pure or contained less than 1.

#### Identification of protein markers

We evaluated the informatics peaks with top importance for the predictive models. Based on the feature importance list (eTable 1), we applied LIFT technology (Bruker Daltonics) to identify the informative peaks whose m/z value was lower than m/z 4000, and the spectral intensity was relatively high enough for the subsequent TOF/TOF process. In the list of peak candidates for S. aureus, an m/z value of 3006 matched the selection criteria. The details of LIFT is described in a previous study^2^.

### eTable 1. Top 15 RF importance of MS peaks.

| **Rank** | **MZ** | **R proportion** | **S proportion** | **Difference** | **RF importance** |
| --- | --- | --- | --- | --- | --- |
| (a) *S. aureus* | | | | | |
| 1 | 6593.357 | 0.6937 | 0.4687 | 0.2250 | 0.0259 |
| 2 | 3296.813 | 0.4381 | 0.1939 | 0.2442 | 0.0212 |
| 3 | 2413.887 | 0.2424 | 0.1072 | 0.1352 | 0.0197 |
| 4 | 6552.397 | 0.8136 | 0.6643 | 0.1493 | 0.0152 |
| 5 | 2430.353 | 0.6162 | 0.5066 | 0.1096 | 0.0145 |
| 6 | 3037.908 | 0.5404 | 0.5158 | 0.0246 | 0.0141 |
| 7 | 7420.699 | 0.1256 | 0.264 | 0.1384 | 0.0114 |
| 8 | 3763.634 | 0.2631 | 0.3484 | 0.0853 | 0.0107 |
| 9 | 4497.474 | 0.3266 | 0.4461 | 0.1195 | 0.0105 |
| 10 | 3277.255 | 0.5699 | 0.6132 | 0.0433 | 0.0102 |
| 11 | 7595.253 | 0.1746 | 0.0326 | 0.1420 | 0.0099 |
| 12 | 2877.45 | 0.4007 | 0.2498 | 0.1509 | 0.0094 |
| 13 | 3006.311 | 0.8063 | 0.8346 | 0.0283 | 0.0092 |
| 14 | 2907.865 | 0.2384 | 0.1588 | 0.0796 | 0.0085 |
| 15 | 4641.999 | 0.2106 | 0.0972 | 0.1134 | 0.0085 |
| (b) *A. baumannii* | | | | | |
| 1 | 6160.411 | 0.0316 | 0.4006 | 0.3690 | 0.0721 |
| 2 | 9074.113 | 0.0108 | 0.1803 | 0.1695 | 0.0285 |
| 3 | 6151.368 | 0.4305 | 0.1496 | 0.2809 | 0.0255 |
| 4 | 2919.033 | 0.4879 | 0.2947 | 0.1932 | 0.0173 |
| 5 | 4107.303 | 0.0194 | 0.1694 | 0.1500 | 0.0165 |
| 6 | 7853.416 | 0.2730 | 0.0416 | 0.2314 | 0.0157 |
| 7 | 12287.38 | 0.1683 | 0.0021 | 0.1662 | 0.0109 |
| 8 | 6143.598 | 0.1999 | 0.0559 | 0.1440 | 0.0102 |
| 9 | 12300.19 | 0.1387 | 0.0105 | 0.1282 | 0.0086 |
| 10 | 6329.521 | 0.7450 | 0.6246 | 0.1204 | 0.0072 |
| 11 | 9086.013 | 0.1968 | 0.0345 | 0.1623 | 0.0072 |
| 12 | 4244.316 | 0.7323 | 0.7230 | 0.0093 | 0.0071 |
| 13 | 3674.093 | 0.0258 | 0.1257 | 0.0999 | 0.0068 |
| 14 | 3965.205 | 0.5625 | 0.6221 | 0.0596 | 0.0068 |
| 15 | 5373.209 | 0.0050 | 0.0664 | 0.0614 | 0.0068 |
| (c) *E. faecium* | | | | | |
| 1 | 6661.806 | 0.4053 | 0.1391 | 0.2662 | 0.019 |
| 2 | 6603.880 | 0.8736 | 0.6459 | 0.2277 | 0.0185 |
| 3 | 3300.989 | 0.9222 | 0.7285 | 0.1937 | 0.0177 |
| 4 | 6689.544 | 0.4197 | 0.1832 | 0.2365 | 0.0143 |
| 5 | 3870.819 | 0.1962 | 0.4079 | 0.2117 | 0.0112 |
| 6 | 6343.554 | 0.9722 | 0.8193 | 0.1529 | 0.0108 |
| 7 | 3171.921 | 0.9767 | 0.8315 | 0.1452 | 0.0092 |
| 8 | 2719.417 | 0.1806 | 0.2774 | 0.0968 | 0.0088 |
| 9 | 3645.827 | 0.9540 | 0.7908 | 0.1632 | 0.0086 |
| 10 | 6512.420 | 0.7159 | 0.8581 | 0.1422 | 0.0078 |
| 11 | 3255.724 | 0.6944 | 0.8349 | 0.1405 | 0.0073 |
| 12 | 3883.734 | 0.9561 | 0.8412 | 0.1149 | 0.0064 |
| 13 | 5094.334 | 0.4584 | 0.3053 | 0.1531 | 0.0063 |
| 14 | 3688.113 | 0.7852 | 0.7897 | 0.0045 | 0.0063 |
| 15 | 3164.015 | 0.0422 | 0.1787 | 0.1365 | 0.0062 |
| (d) *K*. *pneumoniae* | | | | | |
| 1 | 4520.218 | 0.1565 | 0.0230 | 0.1335 | 0.0244 |
| 2 | 5410.193 | 0.6781 | 0.4975 | 0.1806 | 0.0143 |
| 3 | 5938.689 | 0.6847 | 0.5742 | 0.1105 | 0.0108 |
| 4 | 11878.28 | 0.4481 | 0.3005 | 0.1476 | 0.0107 |
| 5 | 4708.548 | 0.1089 | 0.0403 | 0.0686 | 0.0104 |
| 6 | 5070.876 | 0.2652 | 0.1651 | 0.1001 | 0.0087 |
| 7 | 6153.029 | 0.4789 | 0.3508 | 0.1281 | 0.0068 |
| 8 | 5280.115 | 0.8502 | 0.7916 | 0.0586 | 0.0067 |
| 9 | 2617.024 | 0.5880 | 0.6734 | 0.0854 | 0.0062 |
| 10 | 4155.528 | 0.7309 | 0.6490 | 0.0819 | 0.0062 |
| 11 | 7706.079 | 0.8559 | 0.8361 | 0.0198 | 0.0062 |
| 12 | 8309.833 | 0.6562 | 0.5782 | 0.0780 | 0.0061 |
| 13 | 4570.745 | 0.8879 | 0.9311 | 0.0432 | 0.0059 |
| 14 | 2035.513 | 0.6476 | 0.6825 | 0.0349 | 0.0056 |
| 15 | 5322.208 | 0.2063 | 0.3022 | 0.0959 | 0.0052 |
| (e) *E. aerogenes* | | | | | |
| 1 | 4395.726 | 0.3551 | 0.2494 | 0.1057 | 0.0162 |
| 2 | 3784.287 | 0.5513 | 0.6797 | 0.1284 | 0.0112 |
| 3 | 7359.469 | 0.6672 | 0.7539 | 0.0867 | 0.0098 |
| 4 | 7568.535 | 0.6152 | 0.6381 | 0.0229 | 0.0093 |
| 5 | 7648.238 | 0.9302 | 0.9454 | 0.0152 | 0.0083 |
| 6 | 3679.699 | 0.6226 | 0.7196 | 0.0970 | 0.0080 |
| 7 | 2697.051 | 0.9539 | 0.9511 | 0.0028 | 0.0066 |
| 8 | 2221.819 | 0.5661 | 0.5020 | 0.0641 | 0.0064 |
| 9 | 9139.830 | 0.9346 | 0.9650 | 0.0304 | 0.0063 |
| 10 | 3580.358 | 0.9525 | 0.9641 | 0.0116 | 0.0061 |
| 11 | 3824.498 | 0.8975 | 0.9030 | 0.0055 | 0.0061 |
| 12 | 2856.061 | 0.8737 | 0.8794 | 0.0057 | 0.0059 |
| 13 | 7159.944 | 0.9480 | 0.9731 | 0.0251 | 0.0058 |
| 14 | 4570.251 | 0.9287 | 0.9593 | 0.0306 | 0.0057 |
| 15 | 3143.980 | 0.8083 | 0.8191 | 0.0108 | 0.0057 |
| (f) *E. cloacae* | | | | | |
| 1 | 5092.599 | 0.2418 | 0.0956 | 0.1462 | 0.0190 |
| 2 | 5643.628 | 0.6305 | 0.5392 | 0.0913 | 0.0102 |
| 3 | 2820.995 | 0.4559 | 0.3323 | 0.1236 | 0.0083 |
| 4 | 4490.338 | 0.1895 | 0.1361 | 0.0534 | 0.0077 |
| 5 | 5627.055 | 0.3300 | 0.2460 | 0.0840 | 0.0062 |
| 6 | 2852.508 | 0.9109 | 0.8933 | 0.0176 | 0.0058 |
| 7 | 4164.163 | 0.8373 | 0.7590 | 0.0783 | 0.0057 |
| 8 | 5105.856 | 0.2500 | 0.2857 | 0.0357 | 0.0057 |
| 9 | 5663.909 | 0.7923 | 0.7766 | 0.0157 | 0.0057 |
| 10 | 2544.661 | 0.2345 | 0.1500 | 0.0845 | 0.0056 |
| 11 | 3579.762 | 0.8773 | 0.8009 | 0.0764 | 0.0056 |
| 12 | 3845.112 | 0.6073 | 0.4619 | 0.1454 | 0.0052 |
| 13 | 8413.320 | 0.3550 | 0.2302 | 0.1248 | 0.0052 |
| 14 | 6329.332 | 0.9023 | 0.8987 | 0.0036 | 0.0052 |
| 15 | 3621.633 | 0.9509 | 0.9255 | 0.0254 | 0.0050 |

### eTable 2(a). XBugHunter’s potential reduction on anti-MRSA agents in a tertiary medical center for one year. DDD: defined daily dose.

| **Antimicrobial agents** | **Total dose** | **DDD unit (g)** | **Reduced DDD**  **per year** |
| --- | --- | --- | --- |
| **TEICOPLANIN （INJ） 200MG/VIAL** | 468 | 0.4 | 187.2 |
| **VANCOMYCIN HCL （IVF） 1GM/VIAL** | 173 | 2 | 345 |
| **DAPTOMYCIN 500MG/VIAL** | 115 | 0.28 | 32.06 |
| **LINEZOLID INJ,600MG/300ML/BOT** | 9 | 1.2 | 10.2 |

### eTable 2(b). XBugHunter’s potential reduction on anti-MSSA agents in a tertiary medical center for one year. DDD: defined daily dose.

| **Antimicrobial agents** | **Total dose** | **DDD unit (g)** | **Reduced DDD**  **per year** |
| --- | --- | --- | --- |
| **CEFTRIAXONE 1GM/VIAL** | 335 | 2 | 669 |
| **PIPERACILLIN 2GM+TAZOBACTAM 0.25GM）/VIAL（SANDOZ）** | 293 | 14 | 4102 |
| **CEFTAZIDIME 1G/VIAL** | 274 | 4 | 1094 |
| **CEFOPERAZONE SODIUM 500MG + SULBACTAM SODIUM 500MG）/VIAL** | 257 | 4 | 1028 |
| **DICLOXACILLIN SODIUM 250MG/CAP** | 248 | 2 | 495 |
| **CEFADROXIL MONOHYDRATE 500MG/CAP** | 238 | 2 | 475 |
| **CEFEPIME 500MG/VIAL** | 199 | 4 | 796 |
| **TIENAM（IMIPENEM 500MG+CILASTATIN 500MG）/VIAL** | 156 | 2 | 311 |
| **MEROPENEM TRIHYDEATE 250MG/VIAL** | 143 | 3 | 429 |
| **AUGMENTIN 1GM/F.C TAB（CLAVULANIC ACID125MG+AMOXYCILLIN 875MG）** | 120 | 1.5 | 179.25 |
| **CEFAZOLIN SODIUM 1GM/VIAL** | 104 | 3 | 312 |
| **AMOXYCILLIN 1000MG+CLAVULANIC ACID 200MG）/VIAL** | 102 | 3 | 304.5 |
| **CEFTAZIDIME（TATUMCEF）500MG/VIAL** | 82 | 4 | 328 |
| **CEFIXIME 100MG/CAP（PC）** | 49 | 0.4 | 19.6 |
| **FLOMOXEF 1G/VIAL INJ.** | 41 | 2 | 82 |
| **ERTAPENEM 1GM/VIAL** | 38 | 1 | 38 |
| **MEROPENEM TRIHYDEATE 1GM/VIAL** | 34 | 3 | 102 |
| **DORIPENEM 250MG/VIAL** | 33 | 1.5 | 48.75 |
| **MEROPENEM TRIHYDRATE 500MG/VIAL（MEROPENEM SANDOZ,SANDOZ）** | 23 | 3 | 69 |
| **CURAM 625MG/F.C.TAB （CLAVULANIC ACID 125MG +AMOXYCILLIN 500MG）** | 18 | 1.5 | 26.25 |
| **AMPICILLIN 1GM+SULBACTAM 500MG/VIAL** | 16 | 6 | 96 |
| **TAZOCIN（PIPERACILLIN 2GM+TAZOBACTAM 0.25GM）/VIAL** | 14 | 14 | 196 |
| **CEFUROXIME 250MG/TAB** | 13 | 0.5 | 6.5 |
| **AMPICILLIN 500MG/CAP** | 10 | 2 | 20 |
| **CEFUROXIME （SODIUM） 750MG/VIAL** | 8 | 3 | 24 |

### **eFigure 1. Adopting gray zone method to enhance predictive performance.**

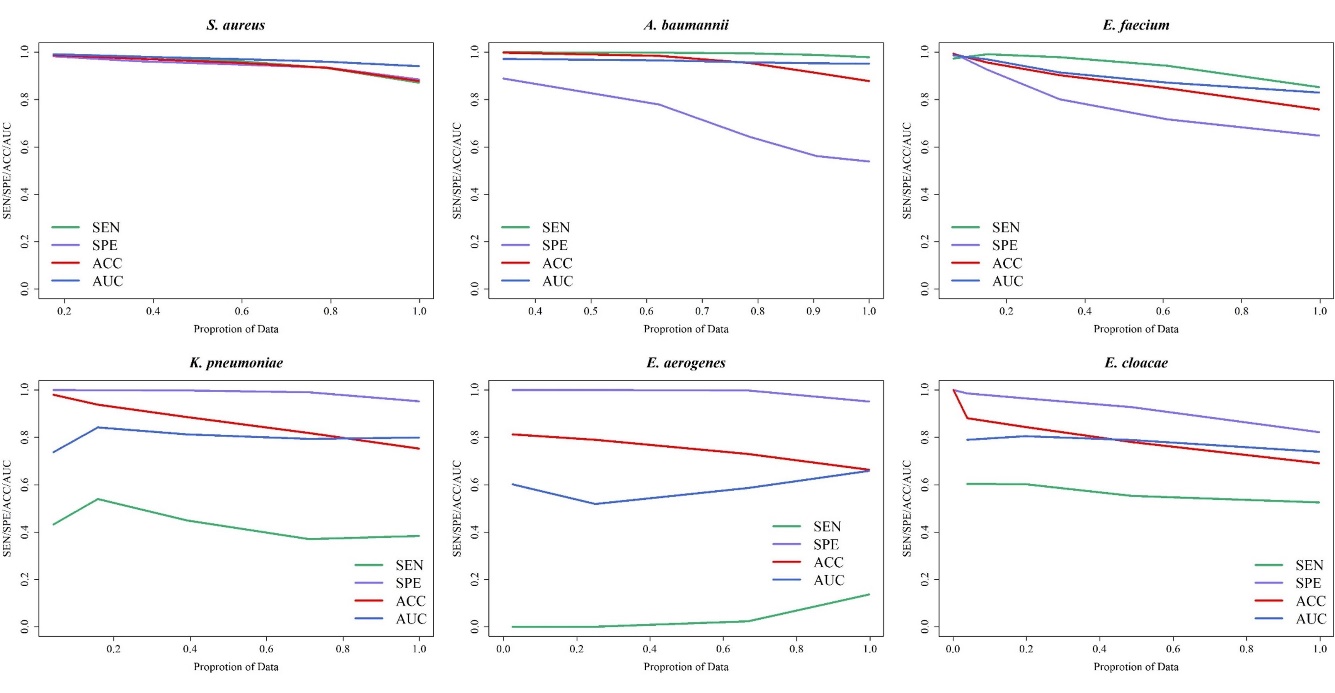
Performance of the random forest under different gray zones (proportions of data). Note that the gray zone was determined around 0.5. In other words, the performance was evaluated for the predicted probabilities which were less than 0.5 − θ or larger than 0.5 + θ. For example, the predicted probabilities which were less than 0.3 were predicted as susceptible while those larger than 0.7 as resistant ones if θ = 0.2. SEN: sensitivity; SPE: specificity; ACC: accuracy; AUC: area under receiver operating characteristic curve.

### eFigure 2. Peptide sequence of *m/z* 3006.

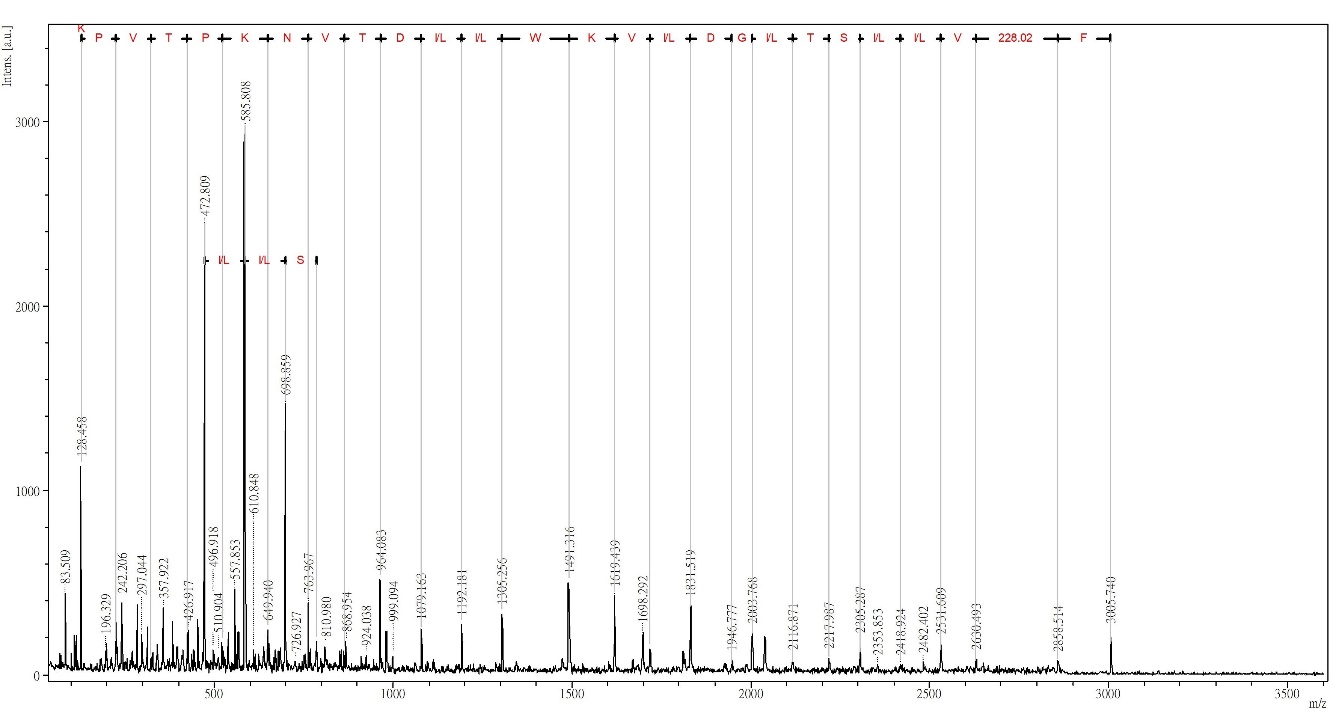

### **eFigure 3(a). Internal validation by using different ML algorithms.**

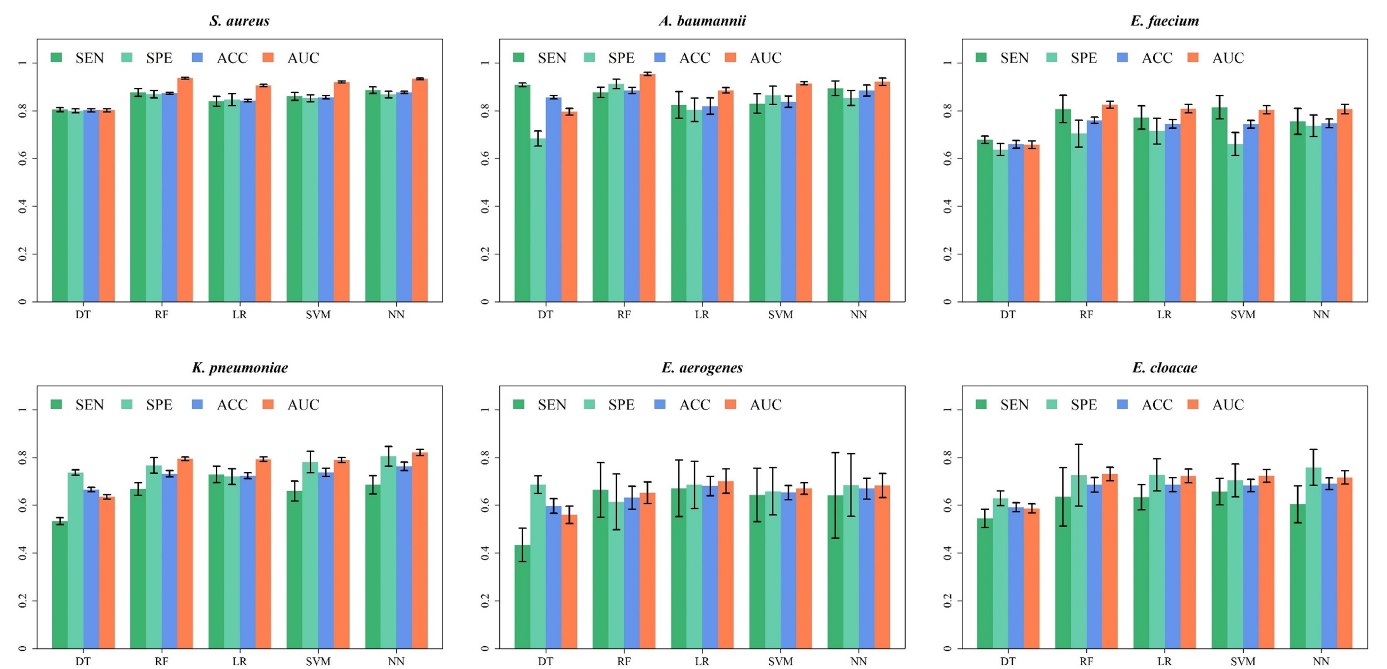

SEN: sensitivity; SPE: specificity; ACC: accuracy; AUC: area under receiver operating characteristic curve. DT: decision tree; RF: random forest; LR: logistic regression; SVM: support vector machine; NN: artificial neural networks.

### eFigure 3(b). Independent testing by using different ML algorithms.

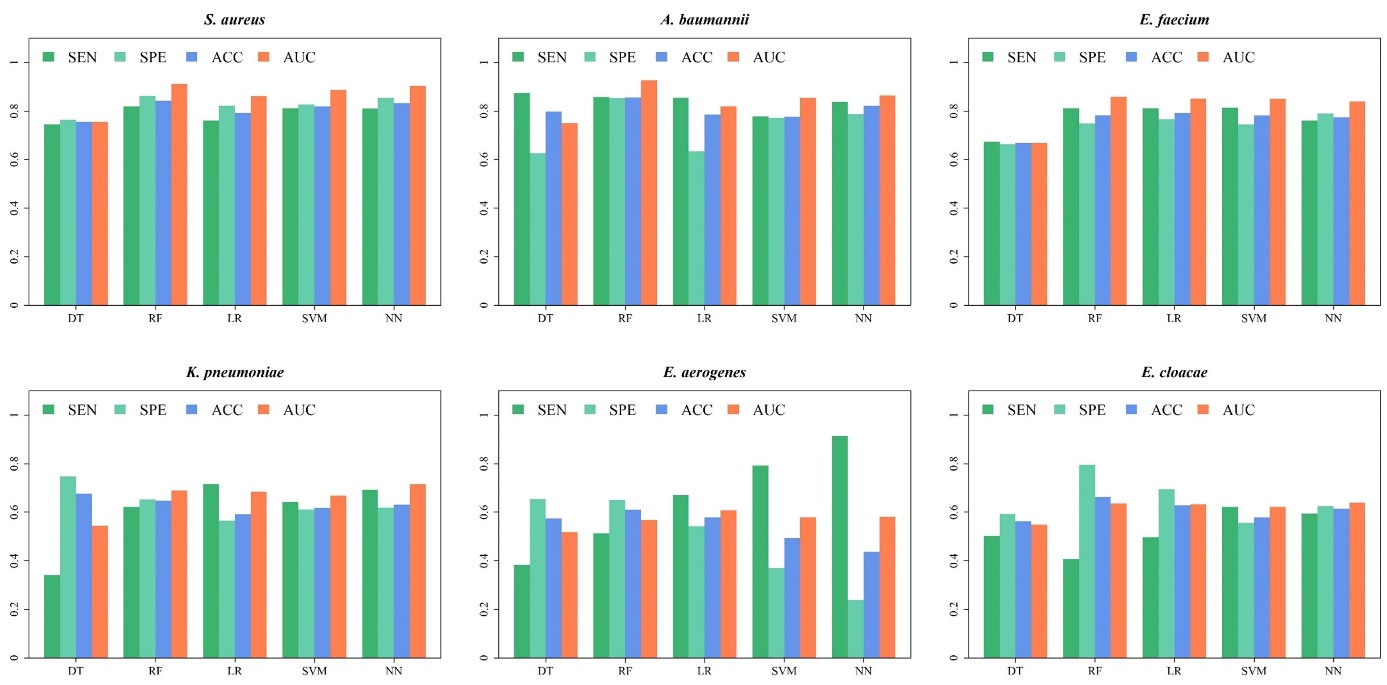

SEN: sensitivity; SPE: specificity; ACC: accuracy; AUC: area under receiver operating characteristic curve. DT: decision tree; RF: random forest; LR: logistic regression; SVM: support vector machine; NN: artificial neural networks.

### eFigure 3(c). Time-wise independent testing by using different ML algorithms.

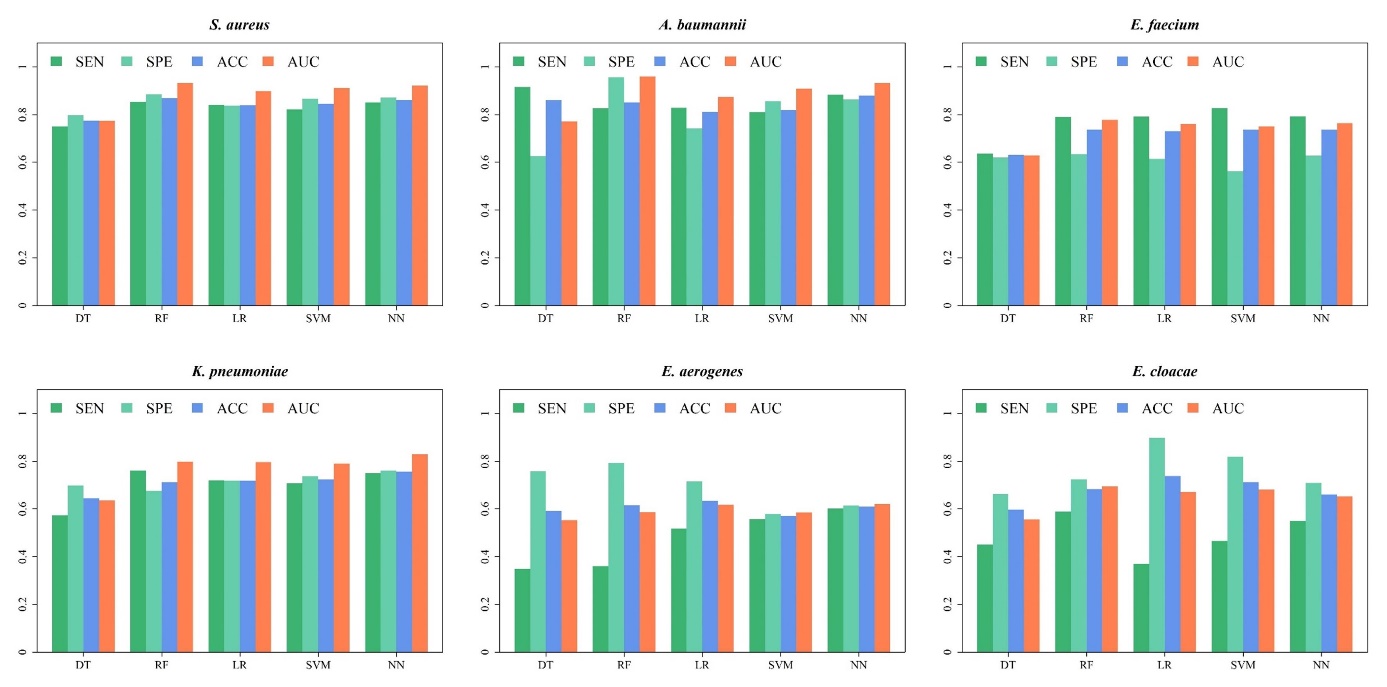

SEN: sensitivity; SPE: specificity; ACC: accuracy; AUC: area under receiver operating characteristic curve. DT: decision tree; RF: random forest; LR: logistic regression; SVM: support vector machine; NN: artificial neural networks.

### eFigure 4(a). Internal validation of different *S. aureus* ML models in a various of specimen types.

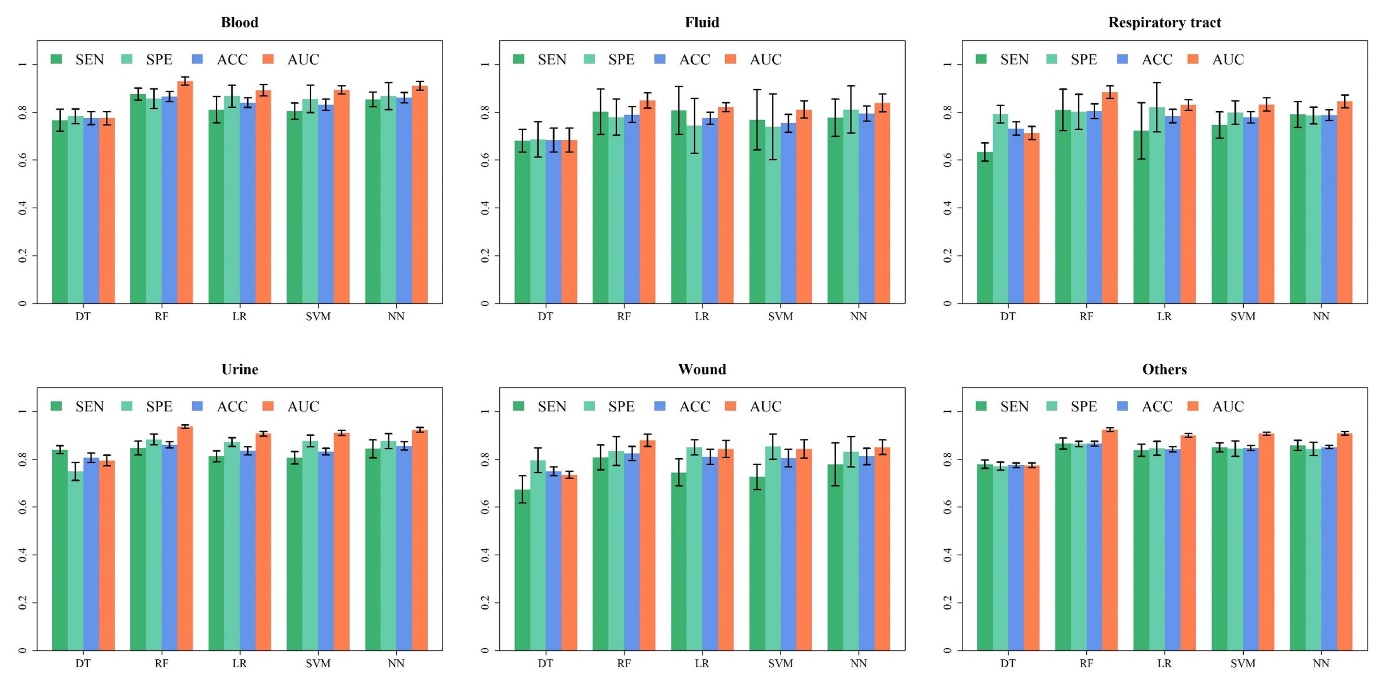

SEN: sensitivity; SPE: specificity; ACC: accuracy; AUC: area under receiver operating characteristic curve. DT: decision tree; RF: random forest; LR: logistic regression; SVM: support vector machine; NN: artificial neural networks.

### eFigure 4(b). Independent testing of different *S. aureus* ML models in a various of specimen types.

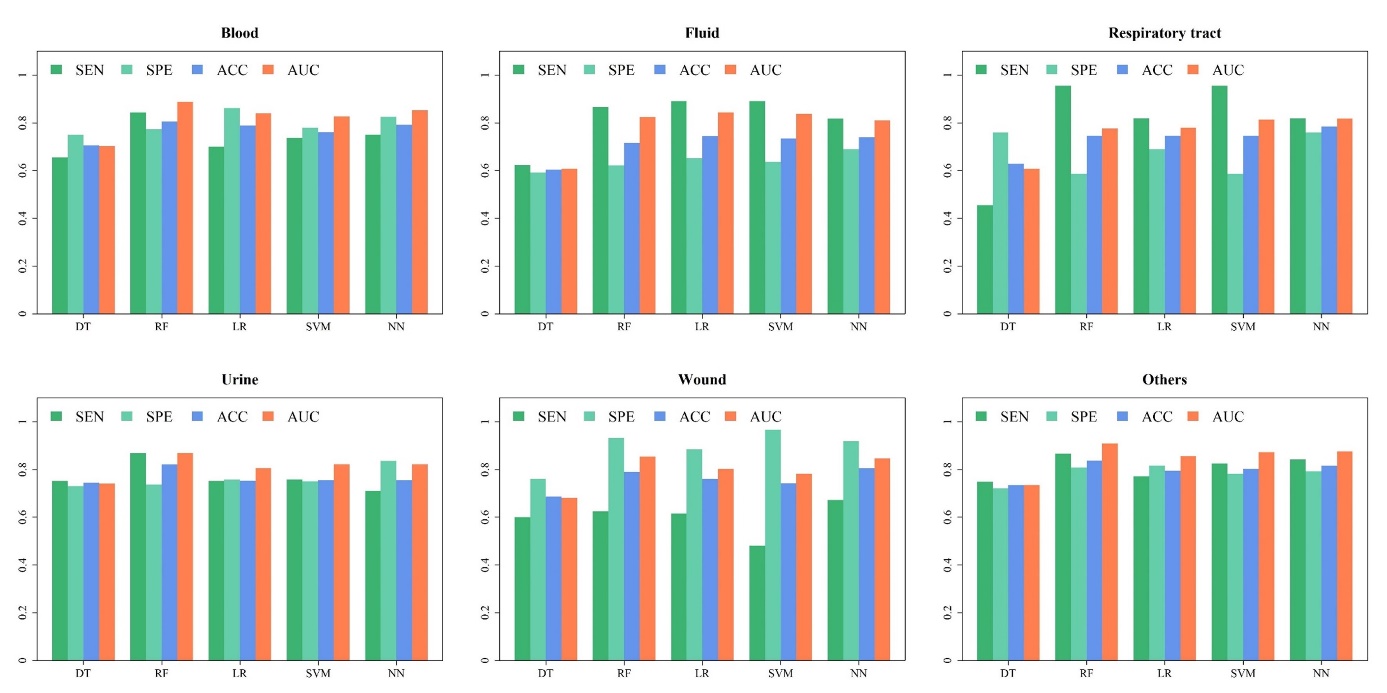

SEN: sensitivity; SPE: specificity; ACC: accuracy; AUC: area under receiver operating characteristic curve. DT: decision tree; RF: random forest; LR: logistic regression; SVM: support vector machine; NN: artificial neural networks.

### eFigure 4(c). Internal validation of different *A. baumannii* ML models in a various of specimen types.

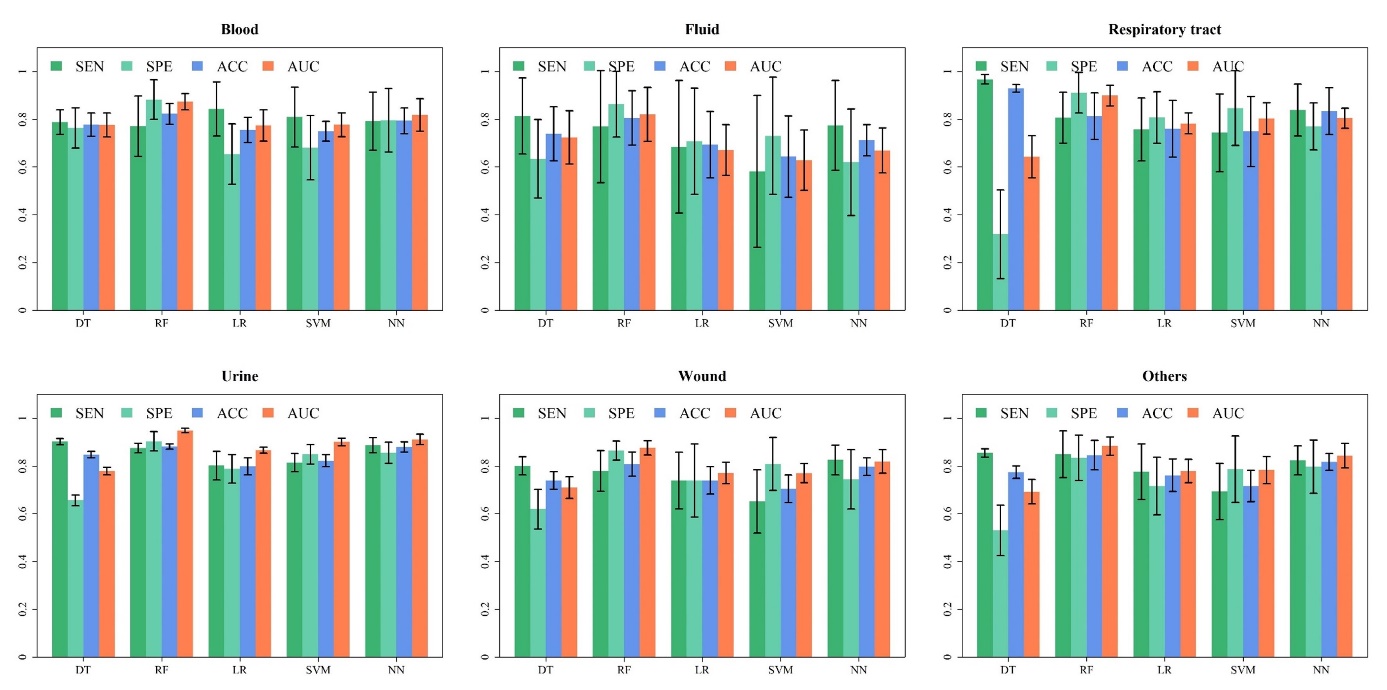

SEN: sensitivity; SPE: specificity; ACC: accuracy; AUC: area under receiver operating characteristic curve. DT: decision tree; RF: random forest; LR: logistic regression; SVM: support vector machine; NN: artificial neural networks.

### eFigure 4(d). Independent testing of different *A. baumannii* ML models in a various of specimen types.

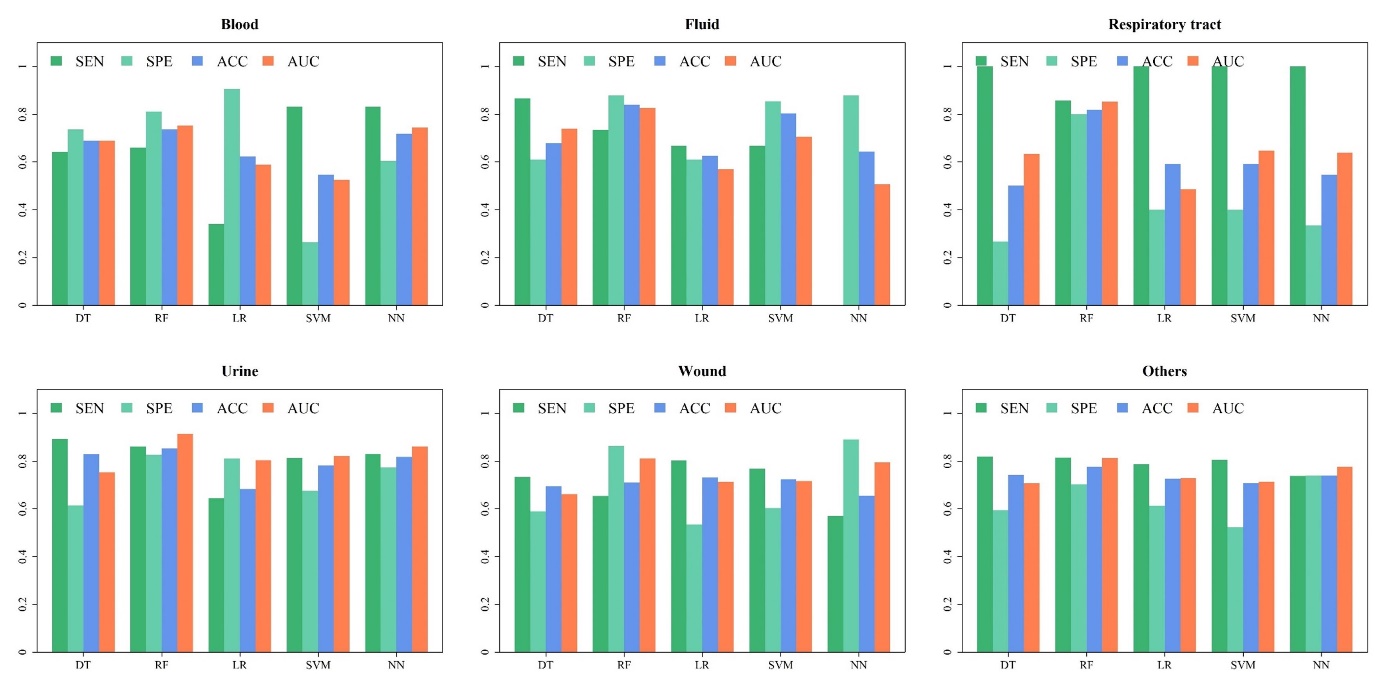

SEN: sensitivity; SPE: specificity; ACC: accuracy; AUC: area under receiver operating characteristic curve. DT: decision tree; RF: random forest; LR: logistic regression; SVM: support vector machine; NN: artificial neural networks.
